# Effectiveness and implementation of “Graduation Approach” livelihood programs with displaced populations: A systematic review

**DOI:** 10.1101/2025.08.01.25332702

**Authors:** Lauren N Yan, Yuwei E Wang, Fred Mubangizi, Colton Parks, Caitlin Whittemore, Nigusu Zelelke

## Abstract

**Background:** Humanitarian actors have widely adopted “Graduation” model programs to support social determinants of health—particularly sustainable livelihoods—among displaced populations. We conducted a systematic review to comprehensively evaluate the evidence regarding Graduation model effectiveness and implementation in displacement contexts. Our objectives were 1) to assess the extent to which Graduation interventions affect food security, employment, living conditions, social wellbeing, poverty, and dependence on humanitarian aid among displaced participants, and 2) to identify barriers and facilitators of implementation in displacement contexts and characterize the model’s feasibility and sustainability in such settings.

**Methods:** We reviewed studies conducted from 2002-2024 in the peer-reviewed and grey literature that explicitly described a “Graduation” intervention (comprising at least three typical components) among displaced adult populations. We excluded studies of economic migrants or host communities and studies with inadequate intervention detail. We searched eight databases (EconLit, Academic Search Ultimate, Family & Society Studies Worldwide, APA PsycINFO, PubMed, Web of Science, Global Health, and the Cochrane Library; updated October 2024) and 11 implementing agencies’ websites (updated April 2024). We assessed study quality using the Mixed Methods Appraisal Tool for systematic mixed-studies reviews. We screened all articles, reviewed full-text studies, and assessed record quality before narratively synthesizing findings.

**Results:** Of 575 screened records, five studies in the grey literature met inclusion criteria (total *N*=28,873 participants). These included randomized trials, program reports, and qualitative studies conducted in Uganda, Mozambique, Colombia, and Costa Rica. Food security outcomes were generally good, but few studies provided quantitative or well-specified estimates. Few employment outcomes were reported; most studies reported improved living conditions. Social wellbeing and poverty outcomes were heterogeneous, with substantial measurement limitations that precluded between-study comparisons. Dependency on humanitarian aid was not directly assessed and mental health outcomes were rarely or vaguely reported. Barriers and facilitators of program implementation were complex, suggesting the importance of comprehensive, community-level support. Program feasibility was best supported by flexible implementation, while sustainability was sensitive to factors such as land tenure and market inter-reliance.

**Discussion:** Most reviewed studies had shortcomings in terms of research rigor and provided insufficient detail to fully assess quality or risk of bias. Findings could not be quantitatively synthesized due to the inconsistency of outcome indicators. While reported findings from these Graduation program evaluations were generally positive, we found little systematic or high-quality evidence for the programs’ effectiveness or implementation in displacement settings. Our review highlights the need to address counterfactuals, long-term outcomes, and implementation concerns in future research. Funding: NIMH grants T32MH103210, T32MH122357, and F31MH136678. Review protocol: PROSPERO CRD42023387899 (May 2023).

## Background

Displacement is a rapidly growing global concern. The number of displaced people worldwide has doubled in the past decade, with estimates exceeding 110 million as of mid-2023 (1). In 2022, 16 new refugees were documented for every one who returned to their country of origin or resettled elsewhere—the highest such ratio ever recorded (1). Internal displacement also reached a record high in 2022, representing a 17% increase in global conflict-related displacements and a 45% increase in disaster-related displacements since 2021 (2).

Employment represents a crucial “social determinant” of overall health (3) and of mental health (4)—yet economic opportunities for displaced people remain elusive, given their tenuous yet protracted circumstances. By the end of 2022, 67% of all refugees remained in protracted situations in low- and middle-income countries (LMICs), having lived in exile or displacement for at least five years; in general, displaced people remain in their hosting communities for an average of 10 years (1,3). While “durable solutions” involving repatriation or resettlement in a third country intend to “[ease] pressure on host countries,” only a small proportion of refugees ever benefit from such solutions (1). Another type of “durable solution” is integration within local hosting communities (1). Sustainable livelihood opportunities for displaced people can facilitate integration by allowing them to provide for their own families, contribute to the local economy, and envision a generative future (6,7)—in turn impacting their comprehensive health and wellbeing.

The Graduation Approach—a multi-faceted livelihood intervention—is one strategy for supporting multiple social determinants of health by developing sustainable livelihoods, which could be a promising approach in LMIC displacement contexts. Introduced by the global non-governmental organization (NGO) BRAC in 2002 as the “Targeting the Ultra-Poor Programme,” this approach aimed to address the holistic needs of an “ultra-poor” subset of the global extreme poor who lived on less than $1.90 per day (8). The original program involved six core program components or “pillars”: 1) life skills and technical training, including weekly home visits or classes; 2) supporting social integration to develop participants’ social networks; 3) providing or facilitating access to health services; 4) teaching financial literacy or establishing savings accounts; 5) short-term consumption support or cash transfers to cover participants’ basic expenses; and 6) a larger-scale asset transfer, e.g., a lump sum of cash or in-kind support to establish a business (9). The Graduation Approach has since gained global recognition as an effective, comprehensive strategy for alleviating severe poverty and its multifaceted health consequences. Six randomized controlled trials conducted in six LMICs concluded that Graduation programs significantly promoted participants’ consumption, self-employment income, and psychosocial status, and that these gains were maintained for at least a year after the intervention in most circumstances (10).

In the past decade, the Graduation Approach has also drawn the interest of major humanitarian actors as they considered strategies to promote displaced people’s livelihoods. In 2014, the United Nations High Commissioner for Refugees (UNHCR) began piloting the Graduation model in humanitarian contexts in Egypt and Costa Rica; these were the first documented efforts to implement the model in displacement settings (12). Early efforts to tailor the Graduation Approach for displacement contexts introduced additional elements such as anti-discrimination campaigns, legal rights education, and psychosocial counseling in an effort to “[empower] refugees and [equip] them with tools for success in their host country” (9). By 2018, 75% of all Graduation programs reported working in “fragile or conflict-affected countries,” and an estimated 16% of all program participants were refugees or internally displaced people (IDPs) (13).

Although Graduation Approach programs are increasingly implemented in displacement settings, their feasibility and effectiveness in such contexts has not been rigorously reviewed. This is concerning, as many features of displacement contexts complicate implementation in ways that could threaten the interventions’ effectiveness. For example, refugees who flee their countries of origin often struggle to acquire formal documentation in host countries, which limits their prospects for economic and social integration (14,15). Displaced communities are often heterogeneous and transient (16,17), introducing challenges for developing appropriate intervention content and following up consistently with participants. Organizations such as World Vision—with which several coauthors are affiliated, and which implements Graduation programs in humanitarian settings—have encountered these challenges firsthand. In one World Vision project site, half of the refugee participants unexpectedly repatriated midway through the program; staff also testified that 3-12 months’ preparation time was needed to properly address the contextual and logistical challenges inherent to humanitarian contexts.

There is a clear need to review and consolidate the evidence base around the Graduation Approach in displacement contexts. To address this gap, we conducted a systematic review to explore all publicly available evidence about the Graduation model’s effectiveness and implementation in humanitarian settings. Specifically, we addressed the following questions:

***1. Graduation outcomes:*** *To what extent do Graduation interventions affect food security, employment, living conditions, social wellbeing, poverty, and dependence on humanitarian aid among displaced participants?*
***2. Graduation implementation:*** *What barriers and facilitators are relevant to program implementation in displacement contexts? How feasible and sustainable is the model’s use in such contexts?*

## Methods

We conducted this study according to the 2020 PRISMA guidelines for systematic reviews (18). Our protocol was registered in the National Institute for Health Research PROSPERO registry in May 2023 (CRD42023387899) (19). We used the PICO(S) (Population, Intervention, Comparison, Outcomes, and Study Type) framework to formulate the eligibility criteria for this study (20).

### Study eligibility

We included studies focusing on displaced populations, including refugees, asylum-seekers, forcibly displaced persons, or other IDPs. We considered studies whose participants were at least 18 years old, and who were willing and able to engage in a livelihood activity. We excluded studies focusing on economic migrants or persons living only in host communities.

We included studies that explicitly described the intervention as the “Graduation approach” or “Ultra-Poor Graduation model,” or that directly referenced BRAC’s “Targeting the Ultra-Poor Programme” as a conceptual template. We only considered programs that included at least three of the following four elements: 1) an asset transfer, 2) consumption support, 3) savings support, and 4) a coaching intervention. We chose these four elements after conducting preliminary scoping reviews, as our findings suggested that most Graduation programs include a combination of these four “pillars” rather than BRAC’s original six core components (9). We excluded studies of programs with fewer than three components; studies that did not explicitly reference the “Graduation approach” or BRAC’s model; and studies that gave inadequate detail for us to determine what the program entailed.

Only quantitative studies were required to have explicitly defined comparator groups, which we defined flexibly (e.g., similar populations not receiving the specified intervention, participants’ own baseline measures, or formally randomized control groups). We imposed no restrictions regarding comparison groups for qualitative studies.

Our primary outcomes of interest were *food security*, *employment, living conditions, social wellbeing, poverty*, *dependence on humanitarian aid,* and *mental health*, as measured at the end of program participation or at later follow-up. We intentionally applied broad definitions of these primary outcome domains, considering the substantial heterogeneity of reported outcomes we noted in an early informal scoping review. As such, we did not exclude any relevant studies based on specific reported measures of effect. Secondary outcomes included *barriers* and *facilitators* (which are factors that hinder or help program implementation, respectively), and programmatic *feasibility* and *sustainability* with displaced populations. *Feasibility* is an intervention’s relevance, suitability, or practicality—typically most salient in early implementation—while *sustainability* refers to an intervention’s maintenance over time, integration into standard practice, and sustained routine use—typically most salient in later implementation stages (21). We extracted data for all compatible measures at all available timepoints (as applicable) from each study.

We excluded documents that were summaries or alternate publications of included studies; purely theoretical articles; articles summarizing evidence from multiple countries; program promotional materials; and articles not describing person-level results. We placed no further restrictions on the types of studies we considered for inclusion.

### Search strategy

We searched eight databases (EconLit, Academic Search Ultimate, Family & Society Studies Worldwide, APA PsycINFO, PubMed, Web of Science, Global Health, and the Cochrane Library). We formulated a search strategy for PubMed (see Table 1) and subsequently adapted it for each database. Our complete search strategies are provided in Supplemental Table 1.

**Table 1.**
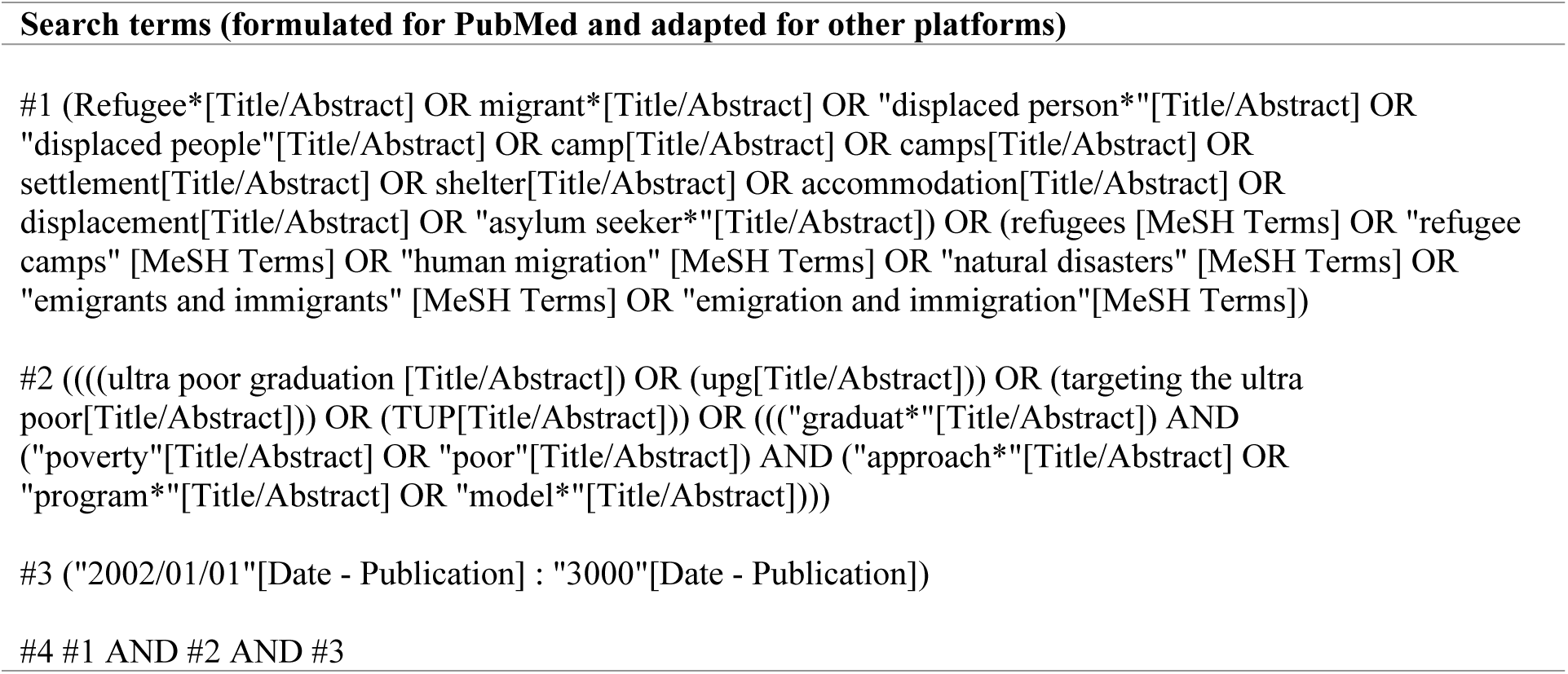
Database search strategy.

| Search terms (formulated for PubMed and adapted for other platforms) |
| --- |
| #1 (Refugee*[Title/Abstract] OR migrant*[Title/Abstract] OR "displaced person*" [Title/Abstract] OR "displaced people" [Title/Abstract] OR camp [Title/Abstract] OR camps [Title/Abstract] OR settlement [Title/Abstract] OR shelter [Title/Abstract] OR accommodation [Title/Abstract] OR displacement [Title/Abstract] OR "asylum seeker*" [Title/Abstract]) OR (refugees [MeSH Terms] OR "refugee camps" [MeSH Terms] OR "human migration" [MeSH Terms] OR "natural disasters" [MeSH Terms] OR "emigrants and immigrants" [MeSH Terms] OR "emigration and immigration" [MeSH Terms])<br>#2 (((ultra poor graduation [Title/Abstract]) OR upg [Title/Abstract])) OR (targeting the ultra poor [Title/Abstract]) OR (TUP [Title/Abstract]) OR ((("graduat*" [Title/Abstract]) AND ("poverty" [Title/Abstract] OR "poor" [Title/Abstract]) AND ("approach*" [Title/Abstract] OR "program*" [Title/Abstract] OR "model*" [Title/Abstract])))<br>#3 ("2002/01/01" [Date - Publication] : "3000" [Date - Publication])<br>#4 #1 AND #2 AND #3 |

When reviewing the grey literature, we hand-searched websites of the following key implementing agencies: Innovations for Poverty Action (IPA), Trickle Up, Poverty Alleviation Coalition, BRAC, Concern Worldwide, World Bank Group Partnership for Economic Inclusion (PEI), Mercy Corps, UNHCR, World Vision, Association of Volunteers in International Service (AVSI), and CARE. We also reviewed the reference list of a key overview publication (22). We restricted our searches to documents published in 2002 or later, and to those published in our study team members’ languages of fluency (English, Chinese, and Spanish). We conducted initial literature searches from March-April 2023, with further grey literature searches in April 2024 after learning of more recently published documents relevant for our review. We conducted a final updated database search in October 2024.

### Study selection and data extraction

Two authors (LY and YW) independently screened document titles and abstracts and removed duplicates using the web-based software platform Covidence (23). Studies were included for full-text review if both reviewers agreed upon their eligibility. During full-text review, LY and YW independently reviewed the articles and decided upon their eligibility for inclusion. All discrepancies were resolved through discussion.

LY and YW extracted data independently using identical Covidence templates with distinct columns for each outcome domain of interest, before comparing extraction results and resolving them into a final document. We recorded study characteristics (full citation, program name, study type, comparison group type, country of implementation, year(s) of implementation); study context (implementation setting, notable details); participant characteristics (number of participants, type of displacement, targeting or inclusion/exclusion criteria, household characteristics); intervention characteristics (program components, duration and sequencing of components, format of implementation, adaptations made); primary effectiveness outcomes (measures of food security, employment, living conditions, social wellbeing, poverty, and humanitarian dependency); and secondary implementation outcomes (barriers, facilitators, feasibility indicators, and sustainability indicators). Where data were missing or unclear, we recorded them as such in the data extraction template.

We did not pre-specify effect measures of interest for each outcome, due to anticipated heterogeneity between studies and accounting for the mixed-methods nature of this review. To facilitate cross-comparison of outcomes, we used our data extraction template to organize each study’s reported outcomes by domain. We assessed the quality of each study using the corresponding measure from the Mixed Methods Appraisal Tool (MMAT) for systematic mixed studies reviews (24).

### Analysis

We reviewed and compared findings across studies using our data extraction template. We narratively synthesized all data from qualitative and quantitative studies, as the level of anticipated heterogeneity in reported outcomes precluded meta-analysis. Our unit of analysis was the *study*, rather than *publication* (multiple publications sometimes reported on the same participant cohort at a given project site; when referring to a *study,* we have triangulated findings from all available *publications* about a given cohort/project). In Covidence, related publication records were grouped together and indexed as a single *study*.

## Results

### Characteristics of included studies

Out of 575 records screened, five distinct studies (reported in six publications) met inclusion criteria. Figure 1 shows a PRISMA flow chart of the screening and assessment process, specifying the number of records assessed at each step (18). After screening out irrelevant references, most publications failed to meet key inclusion criteria; often, they lacked any measure of participant-level results, or their participants were not displaced. Notably, no studies meeting our inclusion criteria were indexed in databases of peer-reviewed literature.

**Figure 1.**
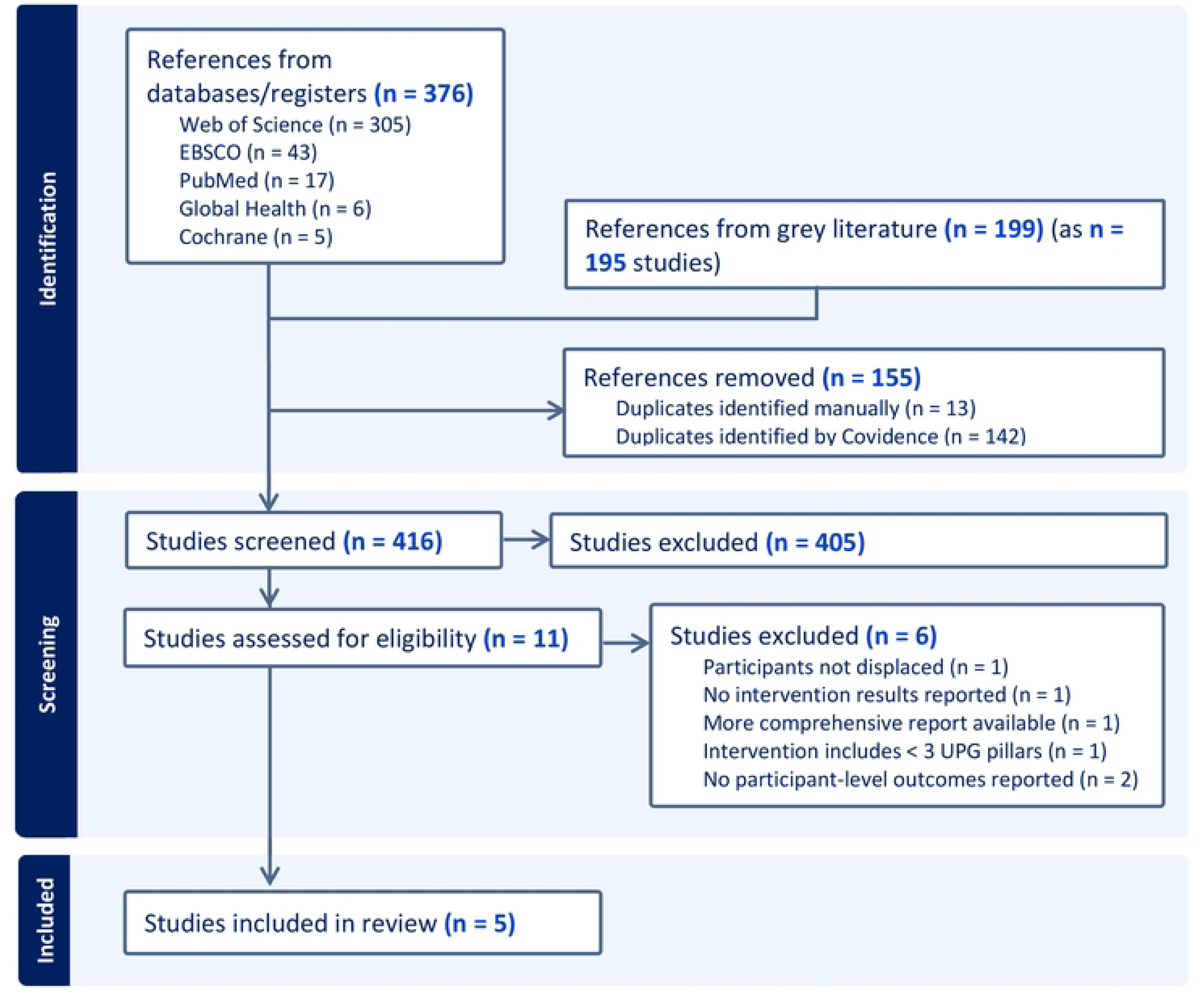
PRISMA flow chart

Several notable studies were excluded at the full text review stage, despite their strong apparent relevance for the study questions. We excluded brief reports and case studies about UNHCR’s pilot projects in Ecuador, which provided insufficient detail about the components (25) and failed to report any results or comparators (15,26). We also excluded the mid-term evaluation of UNHCR’s graduation program in Egypt due to lack of clear implementation of a consumption support pillar (27). Finally, we excluded a case study (28) and a multi-national learning workshop report (29) relating to UNHCR and Self-Help Africa’s Graduation project in Zambia, as neither reported participant outcomes.

The five reviewed studies included two projects in Uganda (30,31), one in Colombia (17,32), one in Mozambique (33), and one in Costa Rica (16). Key characteristics of these publications are described in Table 2. Our final data extraction document after reaching consensus is provided in Supplemental Table 2.

**Table 2.**
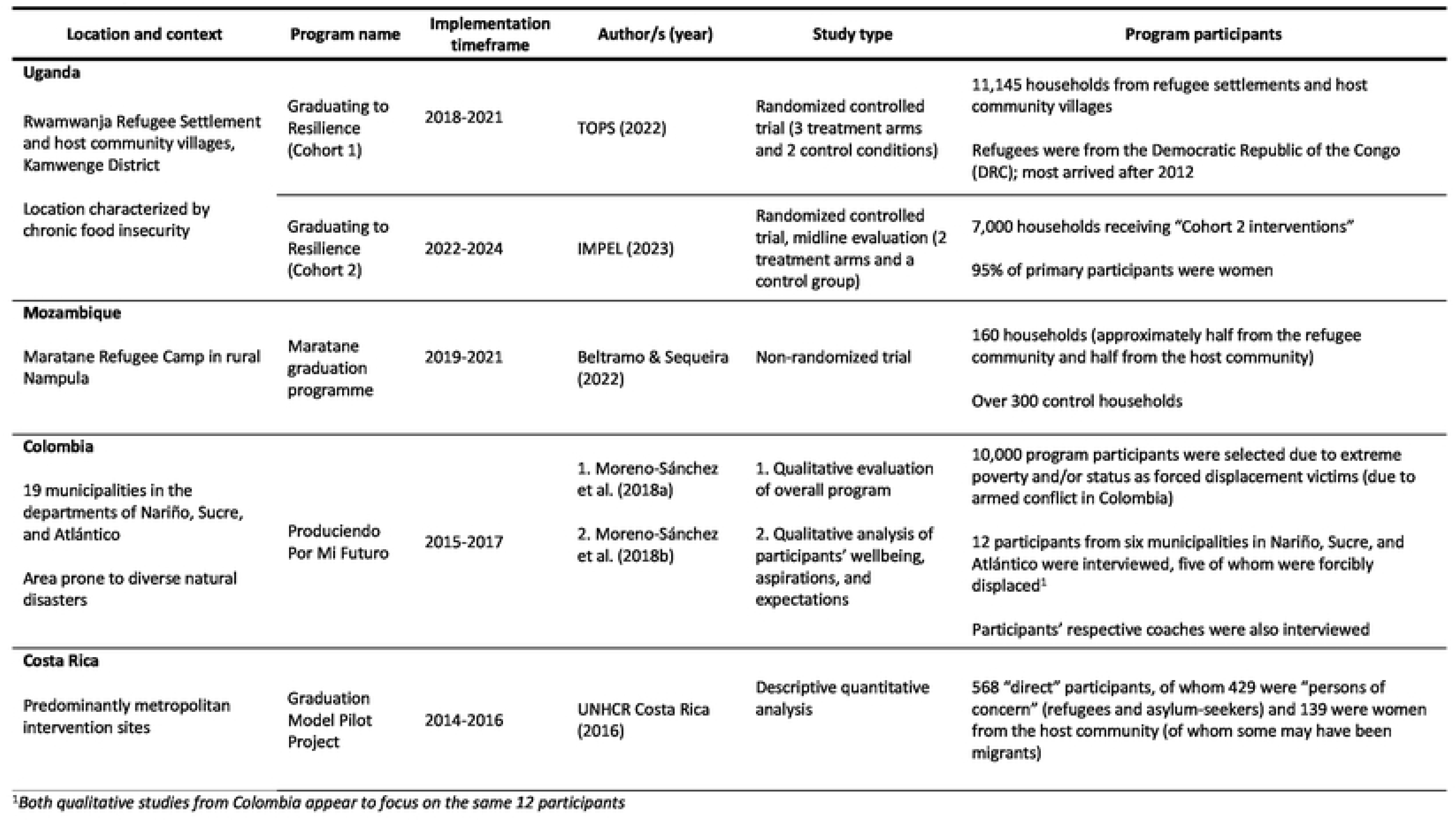
Key characteristics of included studies.

| Location and context | Program name | Implementation timeframe | Author/s (year) | Study type | Program participants |
| --- | --- | --- | --- | --- | --- |
| Uganda |  |  |  |  |  |
| Rwamwanja Refugee Settlement and host community villages, Kamwenge District | Graduating to Resilience (Cohort 1) | 2018-2021 | TOPS (2022) | Randomized controlled trial (3 treatment arms and 2 control conditions) | 11,145 households from refugee settlements and host community villages<br>Refugees were from the Democratic Republic of the Congo (DRC); most arrived after 2012 |
| Location characterized by chronic food insecurity | Graduating to Resilience (Cohort 2) | 2022-2024 | IMPEL (2023) | Randomized controlled trial, midline evaluation (2 treatment arms and a control group) | 7,000 households receiving “Cohort 2 interventions”<br>95% of primary participants were women |
| Mozambique |  |  |  |  |  |
| Maratane Refugee Camp in rural Nampula | Maratane graduation programme | 2019-2021 | Beltramo & Sequeira (2022) | Non-randomized trial | 160 households (approximately half from the refugee community and half from the host community)<br>Over 300 control households |
| Colombia |  |  |  |  |  |
| 19 municipalities in the departments of Nariño, Sucre, and Atlántico | Produciendo Por Mi Futuro | 2015-2017 | 1. Moreno-Sánchez et al. (2018a) | 1. Qualitative evaluation of overall program | 10,000 program participants were selected due to extreme poverty and/or status as forced displacement victims (due to armed conflict in Colombia) |
| Area prone to diverse natural disasters |  |  | 2. Moreno-Sánchez et al. (2018b) | 2. Qualitative analysis of participants’ wellbeing, aspirations, and expectations | 12 participants from six municipalities in Nariño, Sucre, and Atlántico were interviewed, five of whom were forcibly displaced<sup>1</sup><br>Participants’ respective coaches were also interviewed |
| Costa Rica |  |  |  |  |  |
| Predominantly metropolitan intervention sites | Graduation Model Pilot Project | 2014-2016 | UNHCR Costa Rica (2016) | Descriptive quantitative analysis | 568 “direct” participants, of whom 429 were “persons of concern” (refugees and asylum-seekers) and 139 were women from the host community (of whom some may have been migrants) |

<sup>1</sup>Both qualitative studies from Colombia appear to focus on the same 12 participants

The reviewed publications’ quality is described in Table 3 according to the MMAT framework (24). Each publication included at least some elements that were either of poor quality or impossible to assess. The study from Costa Rica did not feature clear research questions, so it was not appropriate to proceed with further quality evaluation using the MMAT (16,24).

**Table 3.**
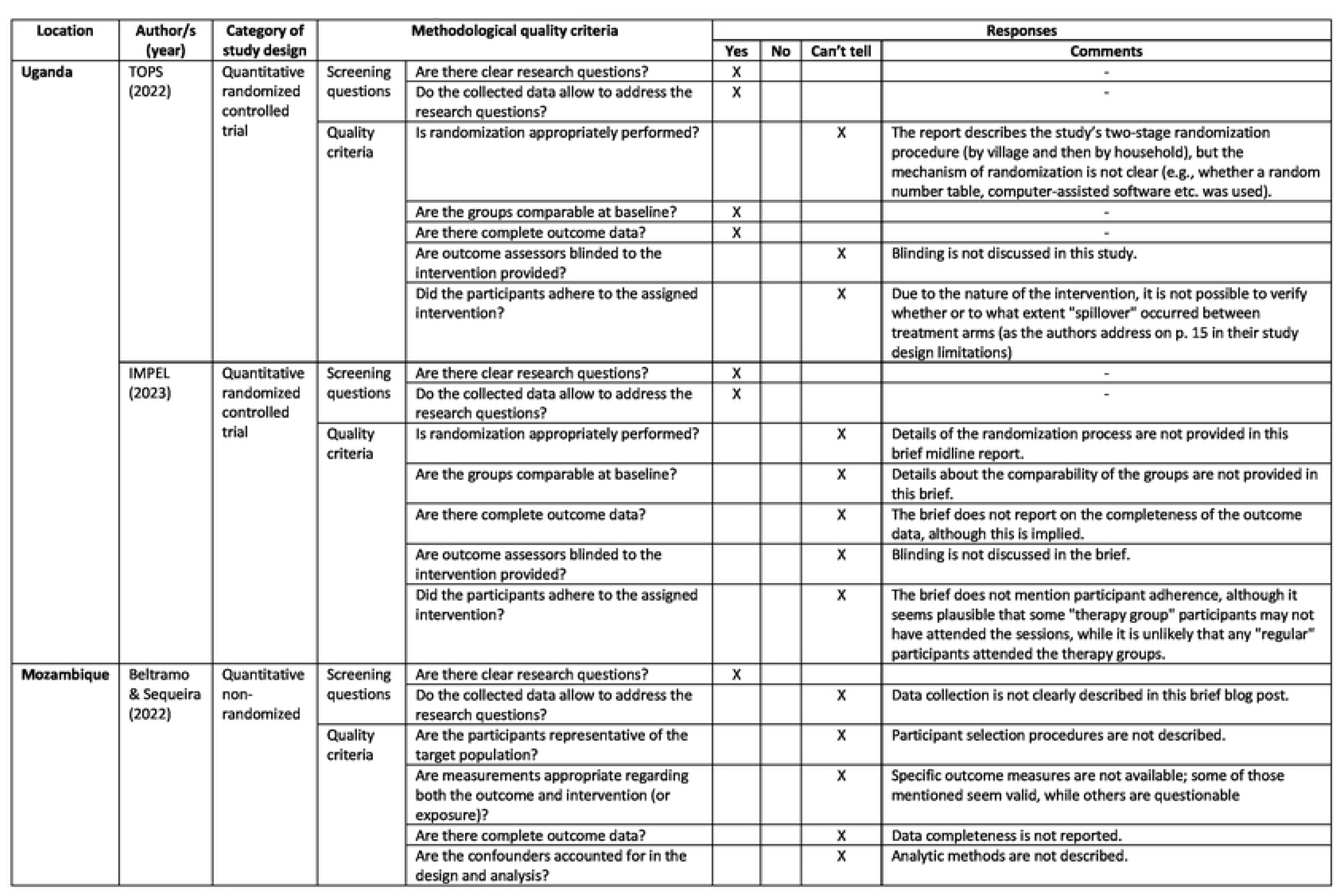
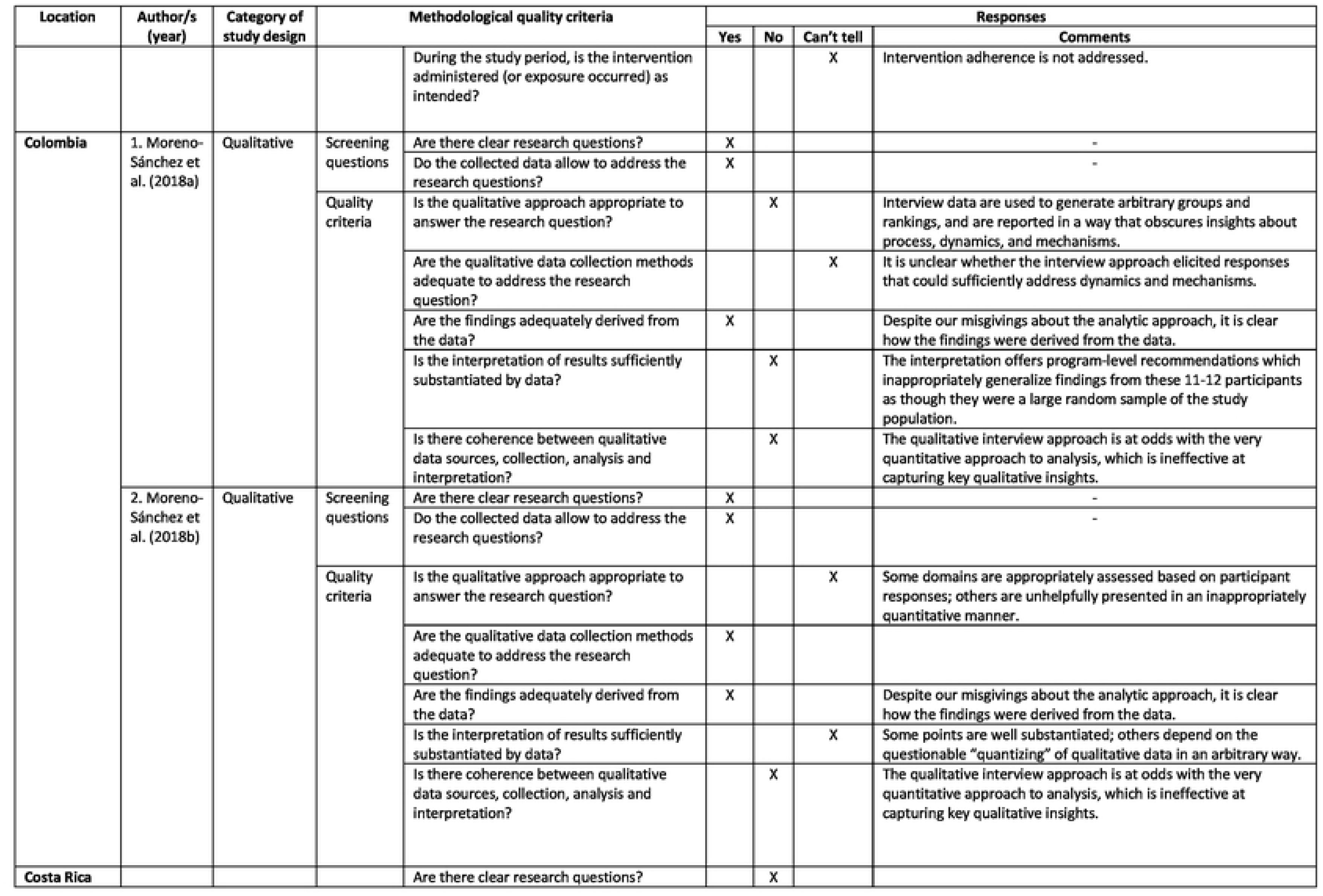
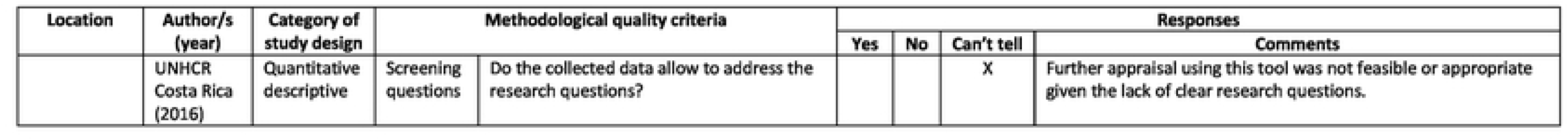
Quality assessment using MMAT.

### Program effectiveness results

#### Food security

Most studies reported improvements in participants’ food security, with broad heterogeneity in outcome measures. The first study in Uganda (representing the first cohort of a continually supported project) reported highly significant increases in food security index scores (comprising measures of food consumption, food insecurity, and children’s length/height-for-age) for all treatment arms relative to controls (30). The second study in Uganda—a midline evaluation of the second cohort, whose participants received adapted intervention components—did not report food security or related measures (31). In Colombia, eight participants (out of only 11) reported in qualitative interviews that they consumed more food, ate more frequent meals, and had more diversified diets during the program (17). In Mozambique, participants reported “lower food insecurity” and had a 40% lower probability of skipping meals than the control group (33). UNHCR’s program report from Costa Rica did not assess food security outcomes. (16)

#### Employment

The reviewed studies reported few employment-related outcomes—except the evaluation of the first Uganda cohort study, which found that program participants in the most comprehensive treatment arm spent significantly more time working than controls (30). In Colombia, seven participants (out of 11) reported diversifying their productive activities during the course of the program, while three others did not; four households who previously engaged in “dependent activities” (e.g., wage employment) increased their time spent on “independent activities” (e.g., self-employment) (17). Three participants also managed to reinvest their profits by the end of the program (17). The Costa Rica study reported endline unemployment rates of 4.6% in the 2014 cohort (versus 37.9% at baseline) and 4.3% in the 2015 cohort (versus 30.4% at baseline) (16). Neither the second-cohort midline study in Uganda (31) nor the study in Mozambique (33) reported employment outcomes.

#### Living conditions and quality of life

Several studies described improvements in participants’ quality of life during the Graduation programs. The first-cohort study in Uganda reported significant improvements in participants’ access to basic drinking water and in handwashing with soap for the most comprehensive treatment arm (with significant improvements in handwashing only for the other treatment arms) relative to controls (30). In Colombia, six participants (out of 11) reported making refurbishments, extensions, or renovations to their houses during the timeframe of the project (17). In Costa Rica, 91.2% of displaced respondents with complete case data finished the project with a valid form of refugee documentation (16). In Mozambique, over 10% of participants bought or constructed a new house, and over 10% invested in electrical grid connections (33). The second-cohort study in Uganda did not describe specific living conditions or quality-of-life improvements (31).

#### Social wellbeing

Studies reported various improvements in participants’ social wellbeing according to heterogeneous assessment measures. The first-cohort study in Uganda reported a highly significant increase in participants’ subjective well-being for all treatment arms relative to controls, measured as an index of participants’ Kessler 6 scores (34) and overall life satisfaction (30). In the second-cohort study in Uganda, host community participants receiving the full intervention (modified to include a group therapy component) scored *lower* on optimism and social wellbeing indices than their counterparts receiving the same support without the therapy component (31). All participants in Colombia reported qualitative improvements in self-esteem, self-value, and self-confidence; some also reported changes in their “aspirations and expectations” (17) or “increases in agency” (32). The study in Costa Rica reported a 9% decrease in the number of participants *not* engaging in “organized groups” over the course of implementation. (16). Of all participants in Costa Rica, 57.9% reported improved emotional wellbeing by the end of the program, with 25.4% reporting no change, 12.3% reporting worse emotional wellbeing, and 4.4% declining to answer (16). In Mozambique, participants reported “significant increases in levels of trust in the other group” (i.e., refugee vs. host community members) by the end of the program, compared to controls (33). The authors also reported that “more participants believ[ed] that both communities [i.e., Mozambiquans and refugees] should be equally prioritized for employment” in circumstances where few jobs are available (33).

#### Poverty

Measures of poverty varied widely between studies, generally finding positive results. In the first-cohort study in Uganda, participants in the all intervention arms had highly significant increases in monthly income, the value of their productive assets, and monthly consumption, relative to controls (30). The second-cohort midline evaluation study in Uganda found that host community participants receiving the most comprehensive treatment (including therapy) scored somewhat lower than participants in the regular intervention on an “economic activity index” (comprising information about labor supply, investment, and intention to expand their enterprise); these differences were driven by disparate levels of investment in income-generating activities (31). The study in Colombia reported qualitative improvements in savings and reinvestment, debt management, spending, and domestic assets for most or all participants (17). Among Costa Rica’s participants with full-case data, 67.5% reported increased income since program baseline, while 18.4% had decreased income, 9.6% maintained their income level, and 4.4% did not answer (16). By the end of the project, 78.9% of participating households reached a monthly income level at least equal to the national minimum wage, while 16.7% did not reach that threshold, and 4.4% did not answer (16). In Mozambique, participants’ household income was 1.3 times greater than controls’ by the end of the program (33). Participants also reported saving an average of $14 per month by the end of the program, while controls reported no savings at all (33).

#### Dependency on humanitarian aid

While dependency on humanitarian aid is often cited as a key problem of protracted displacement that justifies the need for livelihood interventions (11,25,27,35), none of the reviewed studies assessed reliance on aid.

#### Mental health

Few studies assessed mental health outcomes. The second-cohort study in Uganda measured mental health via an “aggregate measure of psychological distress” including the Kessler-10 (34), Patient Health Questionnaire 9 (PHQ-9) (36), and Generalized Anxiety Disorder 7 (GAD-7) questionnaires (31,37). Host community members receiving therapy scored higher on this measure of distress than those in the regular intervention arm; no significant effect was found among refugee participants (31). UNHCR’s study in Costa Rica simply reported that 26.5% of participants or their household members received counseling (11). Other studies did not mention mental health impacts of the Graduation programs.

### Program implementation results

#### Implementation barriers

Barriers to implementation were most extensively addressed in the qualitative research in Colombia. Participants in Colombia experienced multiple livelihood-relevant shocks and obstacles throughout the course of their lives, including illness, employment problems, government corruption, household conflict, regional insecurity and migration shocks, enterprise failure or inability to expand, child school dropout, police involvement with the family, debt, climate shocks, and family deaths (17,32). Participants’ physical isolation from urban centers in Colombia limited the extent of their market integration (17). The authors reported that an “adjustment phase” during program implementation—during which program activities and communication ceased—had also led to a “sense of abandonment and mistrust” that hindered participants’ further progress in the program (32). The first-cohort study in Uganda highlighted infrastructure problems including poor roads, heavy rains, distance between participating households, and COVID-19-related restrictions such as evening curfews that limited staff working time (30). Other studies did not identify implementation barriers.

#### Implementation facilitators

Again, the qualitative research in Colombia described specific facilitators of project implementation in greatest detail. Coaches in Colombia highlighted the importance of participants’ income levels for improving their debt management practices (17). In households where both members of a couple were formal project participants, families achieved higher levels of productive assets (17). The authors concluded that better results tended to occur among participants facing less severe poverty (17). The use of a tablet-based app was viewed favorably in this context, and was more expedient and practical than paper-based material for participants with low literacy (17). Finally, factors thought to improve participants’ education and work prospects were road quality, distance from educational centers, and the quality of available education in the municipality (17). The first-cohort study in Uganda additionally highlighted the cost savings in the treatment arm that included group coaching (rather than one-on one coaching support), which the researchers estimated to be the most cost-effective version of the intervention (30).

#### Feasibility

Studies in Uganda, Colombia, and Costa Rica commented on feasibility. In the first-cohort study in Uganda, key indicators of feasibility were participants’ stability (i.e., likelihood of remaining in their current location rather than repatriating) and their level of interest in the intervention activities (30). In Colombia, the feasibility of establishing livelihood activities was negatively affected by climate shocks, which caused participants to lose livestock and income (17). The authors advocated for flexibly adapting participant recruitment criteria and program components to improve the program’s suitability for diverse participants (17). Finally, because coaches were considered crucial for the program’s success, the authors highlighted the importance of managing coaches’ caseloads to ensure they could maintain a practical workload (17). In Costa Rica, program attrition revealed feasibility issues among certain participant subgroups. A total of 156 participants (27.5%) were dropped from the program due to non-commitment, leaving the country without notice, or losing contact with organization (16). In this context, more refugee participants completed the program (39.6%) than local community participants (22.3%); the authors attributed this difference to local women’s distinct vulnerability statuses and to differences in the intervention (16). A feasible time frame for implementation in Costa Rica was 13-24 months, during which time 72.6% of their “graduated” participants were able to complete the program (16)

#### Sustainability

Only the qualitative research in Colombia provided direct recommendations for program sustainability. Land tenure and environmental shock preparedness were important for sustaining agricultural activities in this context (17). The authors also suggested that the program could better support sustainable enterprises by emphasizing participants’ inter-reliance—ideally, by promoting a range of complementary productive activities to minimize redundancies and competition in restricted markets (17). Finally, the authors suggested that governmental management of Graduation programs as poverty reduction schemes could ensure continuity and consistency of implementation (17).

## Discussion

Although Graduation programs have recently proliferated in displacement contexts (13), only five grey literature studies met our inclusion criteria, indicating severe limitations in the evidence base for Graduation programs’ use in these exceptionally challenging implementation settings. Here, we consider these studies’ implications for future research and practice in the context of previous evidence about the Graduation model.

### Effectiveness

Programs’ reported outcomes were generally positive, but only the “Graduating to Resilience” projects in Uganda utilized randomized controlled trial designs to support causal interpretations of measured improvements (i.e., attributing them to the effect of participating in the programs) (30,31). Specific outcomes and measurement approaches for each outcome domain varied substantially across studies. While most studies lacked the ability to rigorously attribute observed changes to the interventions, their results nevertheless illuminated specific points of interest that implementers and researchers can consider in future work.

While *food security* outcomes were generally good, few studies provided quantitative estimates, and only the first-cohort study in Uganda included well-specified measures (30). Several food security measures are commonly used in humanitarian programming, including the Household Hunger Scale (38), Food Insecurity Experience Scale (FIES) (39), Food Consumption Score (40), or Reduced Coping Strategies Index (41); future Graduation program studies may consider using such measures to enhance comparability. It may be worthwhile to also consider nutrition-related indicators of child growth such as stunting, wasting, and underweight (42), as did the first-cohort study in Uganda using a “food security index” Z-score that combined information from these domains (30). However, while such indices have the advantage of drawing from multiple domains, implementers may have difficulty constructing and interpreting them. From an implementer (and donor) perspective, reporting specific food security-related domains independently may be more interpretable and actionable.

The qualitative research in Colombia highlighted the diversity of factors affecting participants’ food security, including incidental improvements in the food security landscape (e.g., new grocery stores) and other co-existing interventions (17). This raises an important question about how simultaneous supports from multiple projects or sources—sometimes reflecting apparent redundancies in service provision—truly affect program participants. Previous projects have strictly excluded potential participants who already receive comparable support (28). This is a common approach in displacement settings marked by limited short-term funding for lifesaving aid. However, these findings suggest that apparent “redundancies” could potentiate the program’s effectiveness regarding food security (and perhaps other domains as well). This possibility needs to be explored further, and its implications could differ substantially by context (a theme to which we will return frequently). At the very least, these findings caution against excluding participants *a priori* because they receive concurrent support.

Surprisingly few *employment* outcomes were reported, given that the Graduation approach is fundamentally a livelihood intervention. The exception was again the first-cohort study in Uganda, which quantified participants’ total working time (30). The other reviewed studies had limited interpretability due to a lack of nuanced employment indicators (16). For example, some studies focused on the difference between “independence” (e.g., self-employment) and “dependence” (e.g., wage employment) as a key livelihood outcome, with “independence” presumed to be superior (17). However, previous research has demonstrated that this distinction’s meaning is highly context-specific; in some cases, local wage labor represented a more economically attractive opportunity than “independent” employment (43), and “dependency” indicators thus became uninformative or misleading as key outcomes. This finding suggests that “independence” of livelihood activities should not be adopted uncritically or assumed to be a measure of superior employment outcomes.

The reviewed studies also raised questions about whether participants’ enterprise success was due to program elements such as coaching or to external factors (17). Although the question of attributing improvements to the intervention was only addressed rigorously in the first- and second-cohort studies in Uganda, previous work has highlighted the critical importance of addressing counterfactuals. At least one earlier Graduation project documented substantial contextual effects (unrelated to the program) on labor market opportunities (43). In future evaluations, rigorously addressing counterfactuals must become part of standard practice for assessing Graduation programs’ effectiveness.

Four of the five studies reported positive metrics on improved *living conditions* for at least some participants, but again, most (except the first-cohort study in Uganda) did not account for contextual factors that could have affected these outcomes (8). One important insight was that formal documentation is often critical for improving displaced participants’ living conditions; UNHCR’s project in Costa Rica prioritized documentation, recognizing its profound implications for participants’ lives (16). Oher quality-of-life factors that Graduation programs can plausibly impact likely differ across contexts (8,43).

Results for *social wellbeing* demonstrated further measurement limitations, as this construct is difficult to operationalize. The wellbeing index used in the first-cohort study in Uganda included a negative score of the K6 questionnaire (30). However, as this instrument was developed as a screening tool for serious mental illness in the general population (34), not as a wellbeing measure per se, the validity and interpretability of the constructed index are limited. Qualitative research from Colombia presented authors’ and coaches’ conclusions about why wellbeing improved, but did not highlight participants’ own perspectives on this point (32). Participant perspectives will be crucial for future work that seeks to validly assess Graduation programs’ impact on wellbeing. Additionally, quantitative wellbeing metrics such as “group participation” rates in Costa Rica (16) had unclear meaning or practical significance in the absence of further contextual details. Previous research has documented distinct *trajectories* of participants’ holistic wellbeing—trends of improvement, decline, or stagnation over time—and it will be crucial for future studies to interrogate such trends, rather than relying on “snapshots” of a single point in time (45). To accomplish this, detailed subgroup analyses and long-term follow-up studies are needed to clarify how participants may benefit (or decline) through and beyond the duration of the program.

*Poverty* outcomes were highly heterogeneous, as each study used different indicators. Notably, no programs apart from “Graduating to Resilience” in Uganda reported basic indicators such as household consumption, assets, finances, or income/revenue, all of which have been used previously in seminal evaluations of Graduation programs and elsewhere in the economic literature (10). Some studies reported dichotomized or categorical outcome measures, which offer limited information about important poverty-related outcomes such as savings and income (16,17). While the use of dichotomized and categorical indicators often aligns with donor reporting requirements, it seriously limits the inferences that can be drawn from the data. To obtain more informative findings, future studies should consider quantifying impacts more precisely. It is also important to rigorously interrogate the contextual validity of the poverty indicators used in each program. Earlier studies of Graduation programs highlighted the importance of doing so, finding differences in the meaningfulness of savings practices (43), savings group participation (46,47), and quantities saved (10,48) across contexts. Finally, tracking participants’ trajectories of change in poverty measures over time could offer deeper insights about their experience in this domain as well. The one reviewed study that did so found mixed results. In Costa Rica, almost one-third of participants’ poverty measures stagnated or even dropped below baseline levels by the end of the program (16)—a finding which should be flagged for serious follow-up efforts to interrogate potential issues with the program’s effectiveness or implementation. To our knowledge, few studies have followed participants longitudinally after the end of the program, or plan to do so, in displacement settings.

Assessing *dependency on humanitarian aid* presents a significant challenge. Although this is a central issue that motivates Graduation programming in humanitarian contexts, none of the reviewed studies explicitly assessed whether this aim was achieved. Self-reliance (49–52) and resilience (53–57) are stated aims of most UNHCR Graduation programs (25,58,59). This reflects a fundamental departure from traditional Graduation programming, which simply sought to promote entrepreneurial livelihoods in settings of rural poverty—not to enact a shift *away* from existing, reliable subsistence aid. Future projects should explore valid, meaningful metrics to address this issue, engaging in careful qualitative work with participating communities to understand what such a shift would truly entail.

Finally, *mental health* outcomes were only vaguely reported (if mentioned at all) in most studies. It may be advisable for future projects to do so; related programs have found promising results regarding children’s mental health, suggesting that multi-faceted interventions like the Graduation approach could plausibly affect a range of social, emotional, and psychological outcomes at the family level (60). The exception was the second-cohort Uganda project midline report, which focused specifically on adding a group therapy component to the intervention package (31). Again, the project used a composite index score to capture psychological distress; it may be preferable to report the component measures of distress independently, allowing for an assessment of each tool’s contextual validity and its implications for programming. Future studies should consider measuring mental health outcomes with locally adapted and validated measures designed to assess the mental health constructs of interest.

### Implementation

Our findings about Graduation implementation align with broader discussions in the field of implementation science. Implementation is a critical focus of most Graduation literature; we noted that much of the grey literature about Graduation programs focused extensively (often exclusively) on implementation concerns. In particular, a midterm evaluation from Egypt (27) and project reports from Zambia (28,29,35) offered extensive implementation insights. Although we excluded them from the review, they may be informative for Graduation program implementers to consult while working in displacement contexts.

Our findings emphasize the complexity of both *barriers* and *facilitators* of program implementation. Multiple *barriers* were identified, including compounding shocks (environmental, social, or otherwise), obstacles to employment, lack of land tenure, regional insecurity, and distance from urban centers (17,30). Likewise, the studies noted many *facilitators* of program success, sometimes generating comprehensive lists of factors with distinct items, but similar themes, across contexts (17). Family-based social capital was a particularly important facilitator for recovery from shocks (17). This insight draws into question the fundamentally individualistic approach of many Graduation programs, and lends further credence to the finding that additional external support may facilitate greater improvements (17). These findings suggest that more comprehensive, community-level support could be useful if not necessary for Graduation programs’ success.

To address these complex issues, it may be helpful to systematically identify and categorize barriers and facilitators of implementation success by adopting structured determinants frameworks such as the Consolidated Framework for Implementation Research (CFIR) (61,62). Frameworks facilitate a nuanced understanding of barriers and facilitators, helping implementers target strategies to address these challenges (63). Determinants frameworks such as the CFIR not only aid in understanding complex implementation landscapes but also enhance the generalizability and applicability of findings across various contexts.

Our findings suggest that program *feasibility* is best supported by flexible implementation in which programs adapt to participants’ needs, provide additional supports as needed, and pivot strategies in response to shocks (17). The program does not seem practical to implement among communities who move during the course of the program activities, as noted in UNHCR’s Costa Rica program (16), in “Graduating to Resilience” in Uganda (30), and in World Vision’s recent program experience in Zambia. While the time needed to feasibly implement the full program varies by context, it seems unlikely to succeed in less than 13-24 months (16). Funders should seriously consider the implications of such a timeframe, particularly in the context of short humanitarian grant cycles. The program’s multiple components each require substantial time to prepare and implement in these challenging contexts.

Flexible implementation must be supported by carefully documenting and evaluating program adaptations. This need is well-founded in the implementation science literature (64). Tools such as the Framework for Reporting Adaptations and Modifications to Evidence-based Implementation Strategies (FRAME-IS) (65) and the Expert Recommendations for Implementing Change (ERIC) (66) offer valuable methodologies for documenting and assessing contextual adaptations of implementation strategies that enhance the intervention’s feasibility. Comprehensive documentation will be instrumental for rigorously evaluating these strategies, particularly within displacement settings.

Finally, studies suggested that program *sustainability* is sensitive to factors such as land tenure, which is particularly important for many participants’ livelihoods (17). However, land availability is limited in many displacement settings and is likely to present a challenge to the program’s long-term success. Importantly, the Colombia research team also identified the importance of market *inter-reliance* for sustaining positive livelihood outcomes; in contrast, many current Graduation programs present a high risk of creating markets that are both redundant and highly competitive (17). It is crucial to design livelihood strategies that have sustained communal impact, focusing on complementary value chains to promote long-term generative economic activity. Finally, studies suggested that government actors are uniquely well positioned to sustain Graduation-type programs (17). Indeed, Graduation interventions have been integrated into government social safety nets in multiple contexts (15,55,67). This option could powerfully support Graduation programs’ long-term sustainment—although it is unclear whether displaced participants could easily benefit from a nationalized version of the program. Once again, these findings underscore the urgent need for more rigorous evaluation of sustainment strategies in displacement contexts.

Our review highlights a notable gap in research regarding sustainment and longitudinal evaluation of Graduation interventions in displacement contexts. Sustaining Graduation interventions over time requires deep understanding of the interplay between interventions, socio-cultural and community contexts, and organizational factors (68). This understanding begins developing early in the intervention’s lifecycle—even during the planning phase—and develops with ongoing monitoring and collaborative adaptation to evolving contextual needs (69). Addressing this gap requires a committed effort to conducting longitudinal research and developing frameworks that effectively capture long-term outcomes to assess these interventions’ sustainability. Doing so can ensure their continued relevance and impact for addressing the many challenges that displaced populations face.

Graduation programs offer unique opportunities to test and refine existing theories of implementation science in highly dynamic and challenging environments. As such, they demand innovative methodological approaches to evaluation that will be highly relevant for global displacement contexts and for the broader field of implementation science.

### Limitations

Most reviewed studies each had shortcomings in terms of research rigor and provided insufficient detail to fully assess the quality of their work. However, it is important to recognize that none of these documents was intended for peer-reviewed publication. Rather, they provided a full cost-effectiveness report (30), a midline implementation report (31), interim findings clearly intended for more comprehensive write-ups at a later time (33) or provided general overviews for implementation stakeholders (16,17,32). The studies in Uganda (30,31) and Mozambique (33) clearly had high levels of research support, but described their study procedures as suited their document formats (a long report, a brief report, and a blog, respectively). The qualitative studies in Colombia (17,32) provided more transparent descriptions of their study processes, but had shortcomings related to the mismatch of their analytic approach with the nature of their data.

We could not directly assess any of the studies’ risk of reporting bias (e.g., due to missing results), as the level of available detail precluded such an assessment. Furthermore, we could not quantitatively synthesize our findings due to the absolute inconsistency of outcome indicators; no two studies reported *any* outcomes using comparable indicators. Only the studies in Uganda (30,31) assessed statistical or measurement uncertainty. Only the first-cohort Uganda study (30) presented subgroup-specific results for refugee participants (if relevant, i.e., if the participant population included non-displaced host country citizens). We have only reported this study’s combined-sample results, to align with results from other studies that did not distinguish between these groups. Finally, we did not reach out to all program implementers directly to ask about available data.

## Conclusion

Graduation programs have recently drawn strong interest as a valuable strategy for promoting sustainable livelihoods in protracted displacement settings (11,13,70–72). Donors and implementers have high confidence in these programs due to positive early findings (10), which have been widely cited to justify their scale-up in displacement settings (12).

However, our review found relatively limited evidence about effectiveness and implementation for the Graduation approach in displacement settings, with only one rigorous study completed to date (30). More implementation insights are available, but further work is needed to systematically synthesize these findings. This point is especially important—and urgent—for at least two reasons.

First, “traditional” Graduation programs may require substantial adaptation for displacement contexts. Future research needs to account for this heterogeneity of implementation contexts and intervention components. Even traditional Graduation programs contextually adapt components and criteria at each new site (9); as such, assessing program “fidelity” is not feasible, which in turn complicates efforts to evaluate the program’s effectiveness. Future research must face the challenge of rigorously evaluating each program’s effectiveness, using outcome indicators that are both contextually valid and meaningfully comparable to those of other programs. It is critical to address counterfactuals and to conduct detailed subgroup analyses and longitudinal studies to clarify the program’s causal impacts on participants over time, and to determine whether effects are sustained after the end of the program.

Second, current knowledge about implementation factors is difficult for implementers to find or review—and even more challenging to operationalize in their distinct program contexts. Enhanced documentation of adaptations and implementation strategies would be greatly helpful. If this information is publicly available, it can aid implementers in discerning common challenges and opportunities across displacement contexts, and in highlighting important contextual differences. Such documentation could also facilitate development of clearer “standard operating procedures,” which implementers highly value.

The current moment presents a critical opportunity for researchers and implementers to rigorously evaluate Graduation programs in displacement contexts. Longitudinal research that appropriately addresses counterfactuals will be particularly important to inform frameworks that effectively capture long-term outcomes and assess the interventions’ sustainability. The value of assessing Graduation in displacement settings extends beyond immediate programmatic improvement. Such evaluations can inform evidence-based policy and can ultimately facilitate more sustainable and resilient support systems for displaced populations around the world.

## List of Abbreviations

LMICs: Low- and middle-income countries
NGO: Non-governmental organization
UNHCR: United Nations High Commissioner for Refugee
IDPs: Internally displaced persons
IPA: Innovations for Poverty Action
PEI: Partnership for Economic Inclusion (PEI)
AVSI: Association of Volunteers in International Service (AVSI)
MMAT: Mixed Methods Appraisal Tool

## Data availability

All data generated or analyzed during this study are included in this article and its supplementary information files.

## Acknowledgements

The authors gratefully acknowledge Dr. Emmanuel Tumusiime for his early input on conceptualizing this study and for his suggestions for improving the draft manuscript; Dr. Sarah Murray for her advice regarding the scope and approach of the study; and Dr. Christine Bourey for generously supporting the protocol development and overall process of managing this study.

---

**Supplemental Table 1.** Complete search terms for all reviewed databases.

| Database(s) | Search String |
| --- | --- |
| EconLit, Academic Search Ultimate, Family & Society Studies Worldwide, and APA PsycINFO | All searched through EBSCO<br>S1: AB ( Refugee* OR migrant* OR "displaced person*" OR "displaced people" OR camp OR camps OR settlement OR shelter OR accommodation OR displacement OR "asylum seeker*" ) OR SU (refugee OR "refugee camps" OR "human migration" OR "natural disasters" OR "emigrants and immigrants" OR "emigration and immigration") OR TI ( Refugee* OR migrant* OR "displaced person*" OR "displaced people" OR camp OR camps OR settlement OR shelter OR accommodation OR displacement OR "asylum seeker*" )<br>S2: AB ("ultra poor graduation" OR "upg" OR "targeting the ultra poor" OR "TUP" OR (("graduat*" AND ("poverty" OR "poor") AND ("approach*" OR "program*" OR "model*")))) OR TI ("ultra poor graduation" OR "upg" OR "targeting the ultra poor" OR "TUP" OR (("graduat*" AND ("poverty" OR "poor") AND ("approach*" OR "program*" OR "model*"))))<br>S3: S1 AND S2 AND (Published Date: 20020101-20241012) |
| PubMed | #1 (Refugee*[Title/Abstract] OR migrant*[Title/Abstract] OR "displaced person*" [Title/Abstract] OR "displaced people" [Title/Abstract] OR camp [Title/Abstract] OR camps [Title/Abstract] OR settlement [Title/Abstract] OR shelter [Title/Abstract] OR accommodation [Title/Abstract] OR displacement [Title/Abstract] OR "asylum seeker*" [Title/Abstract]) OR (refugees [MeSH Terms] OR "refugee camps" [MeSH Terms] OR "human migration" [MeSH Terms] OR "natural disasters" [MeSH Terms] OR "emigrants and immigrants" [MeSH Terms] OR "emigration and immigration" [MeSH Terms])<br>#2 (((("ultra poor graduation" [Title/Abstract]) OR (upg [Title/Abstract])) OR (targeting the ultra poor [Title/Abstract])) OR (TUP [Title/Abstract])) OR (((("graduat*" [Title/Abstract]) AND ("poverty" [Title/Abstract] OR "poor" [Title/Abstract]) AND ("approach*" [Title/Abstract] OR "program*" [Title/Abstract] OR "model*" [Title/Abstract]))))<br>#3 ("2002/01/01" [Date - Publication] : "3000" [Date - Publication])<br>#4 #1 AND #2 AND #3 |
| Global Health (Ovid) | #1 (Refugee* or migrant* or "displaced person*" or "displaced people" or camp or camps or settlement or shelter or accommodation or displacement or "asylum seeker*").ab,cw,hw,ot,ti. or (refugee or "refugee camps" or "human migration" or "natural disasters" or "emigrants and immigrants" or "emigration and immigration").sh.<br>#2 ("ultra poor graduation" or "upg" or "targeting the ultra poor" or "TUP" or ("graduat*" and ("poverty" or "poor") and ("approach*" or "program*" or "model*"))).ab,cw,hw,ot,ti.<br>#3 1 AND 2 AND (yr= "2002-Current") |
| Web of Science | #1 TS=(Refugee* or migrant* or "displaced person*" or "displaced people" or camp or camps or settlement or shelter or accommodation or displacement or "asylum seeker*") OR TS=(refugees or "refugee camps" or "human migration" or "natural disasters" or "emigrants and immigrants" or "emigration and immigration")<br>#2 TS=("ultra poor graduation" or "upg" or "targeting the ultra poor" or "TUP" or ("graduat*" and ("poverty" or "poor") and ("approach*" or "program*" or "model*")))<br>#3 1 AND 2 AND (publication date: 2002-01-01 to 2024-10-12) |
| Cochrane | ((Refugee* or migrant* or "displaced person*" or "displaced people" or camp or camps or settlement or shelter or accommodation or displacement or "asylum seeker*") OR (refugees or "refugee camps" or "human migration" or "natural disasters" or "emigrants and immigrants" or "emigration and immigration")):ti,ab,kw AND ((("ultra poor graduation" or "upg" or "targeting the ultra poor" or "TUP" or ("graduat*" and ("poverty" or "poor") and ("approach*" or "program*" or "model*")))):ti,ab,kw |

**Supplemental Table 1.**
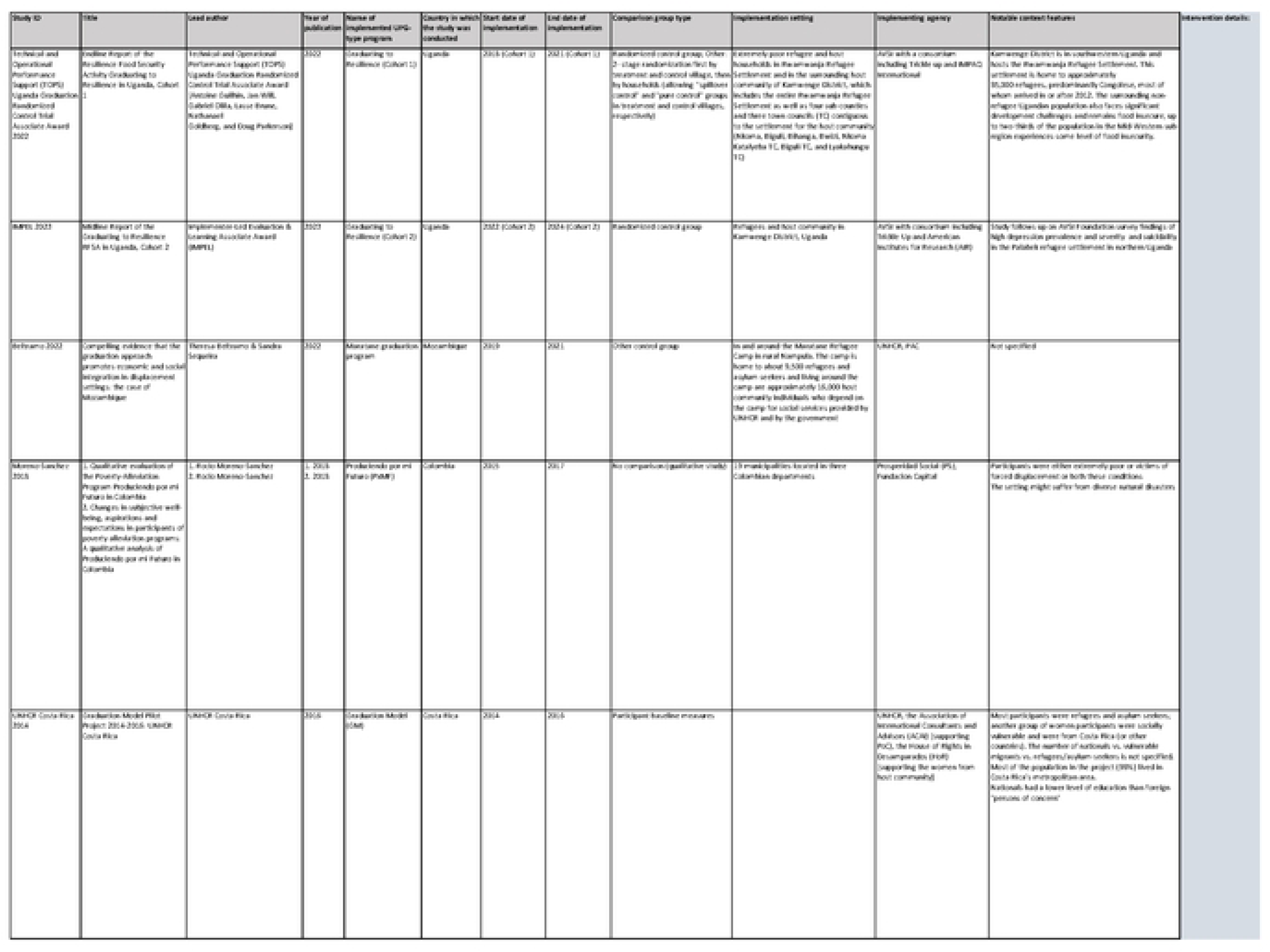
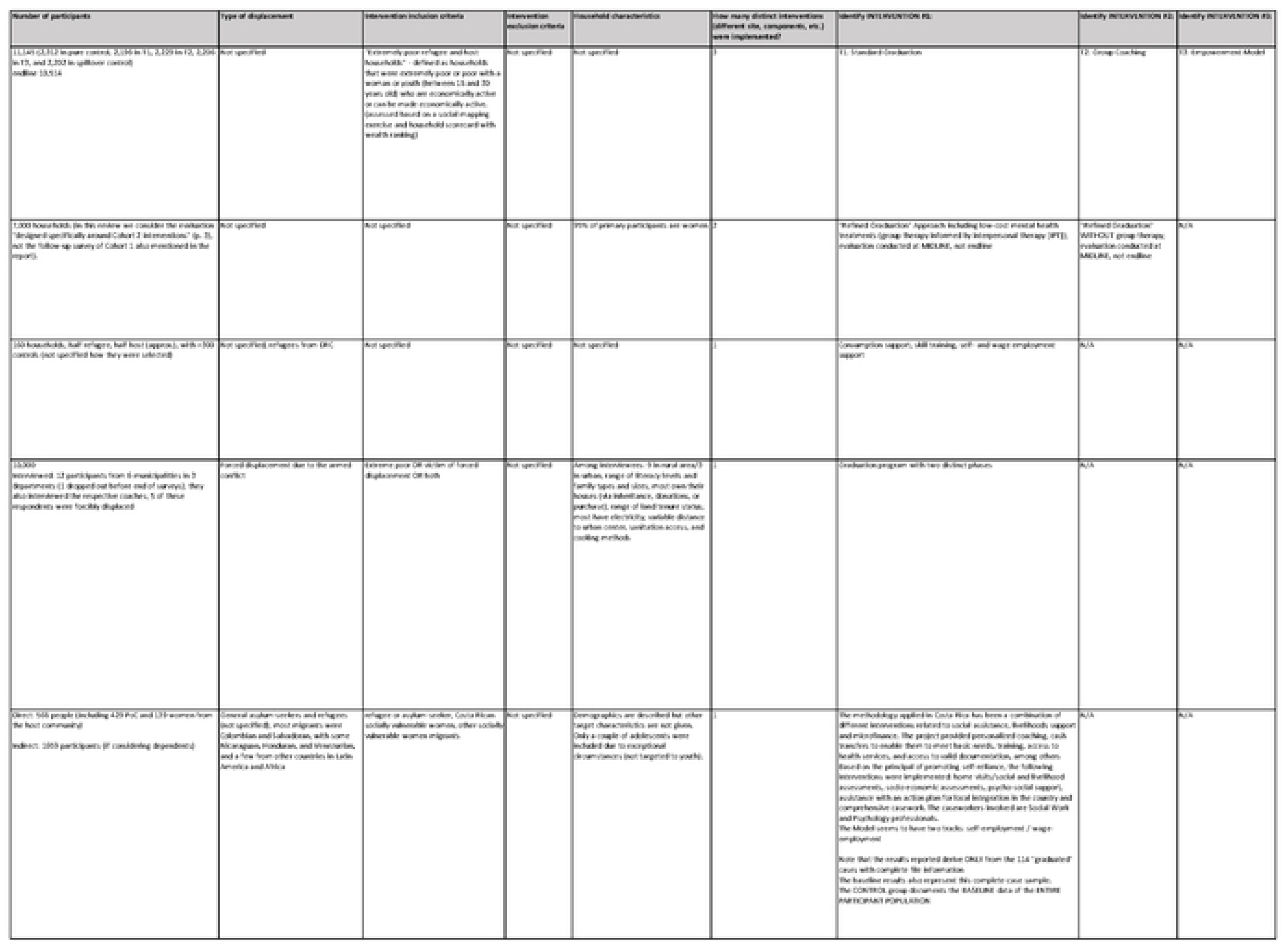
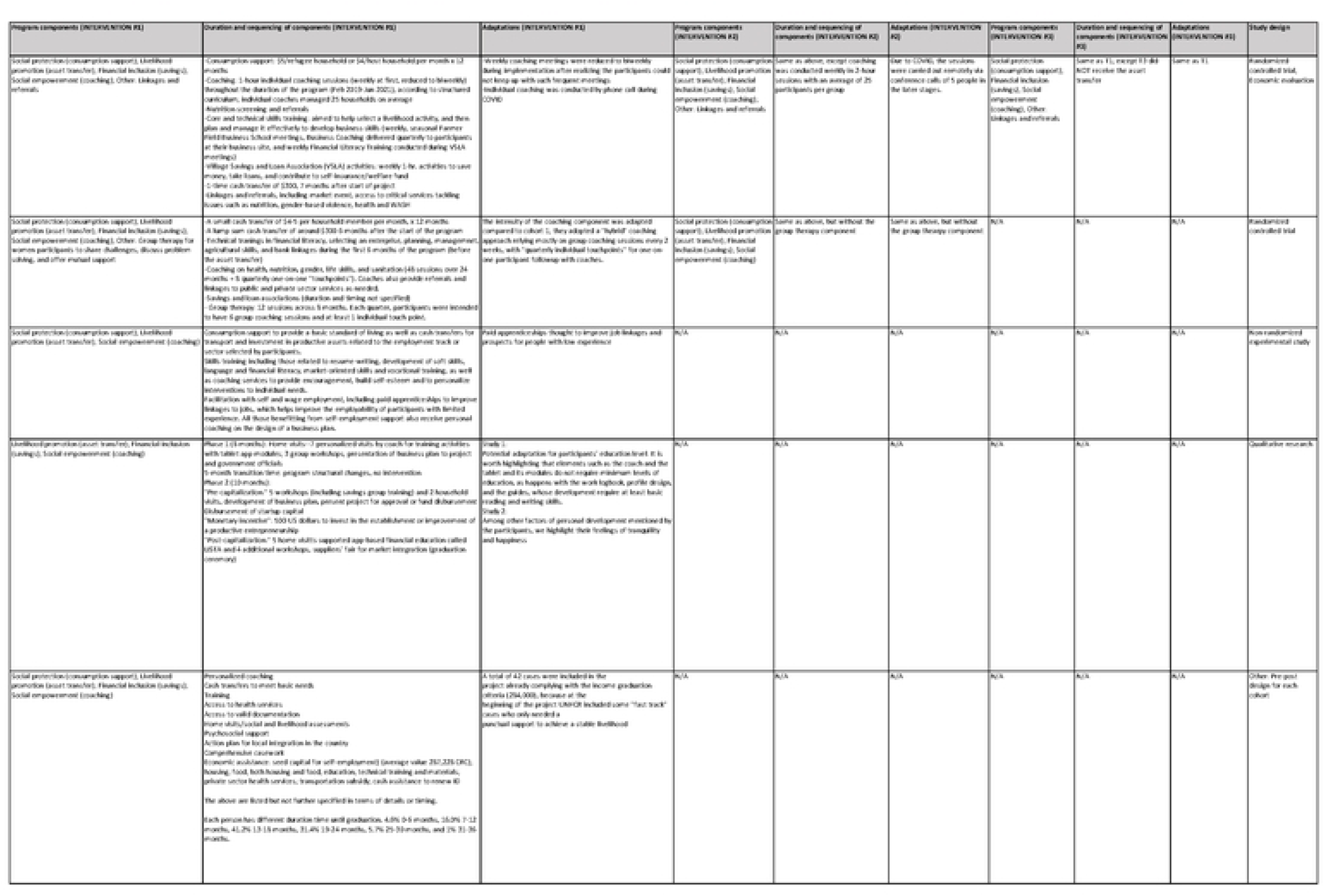
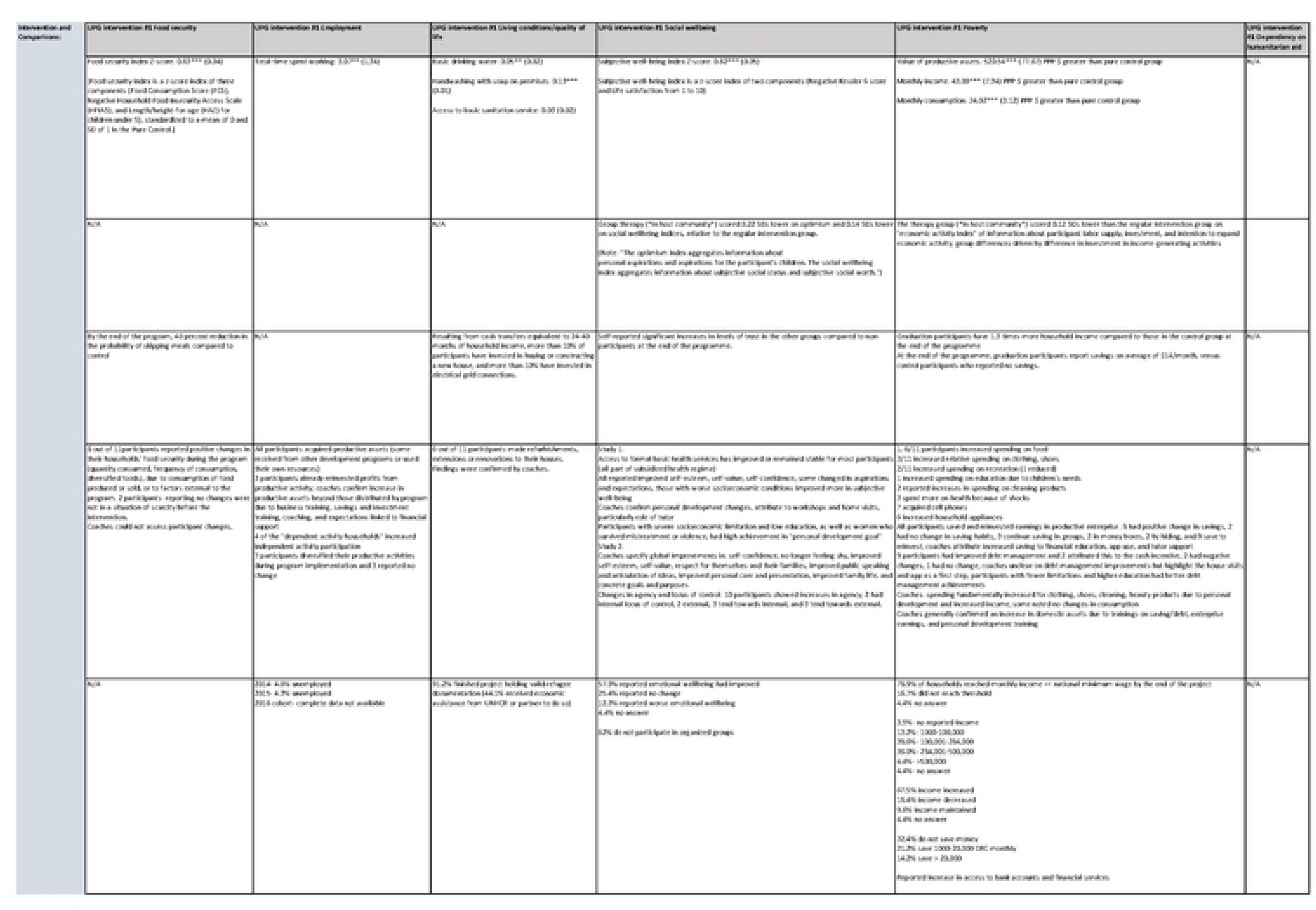
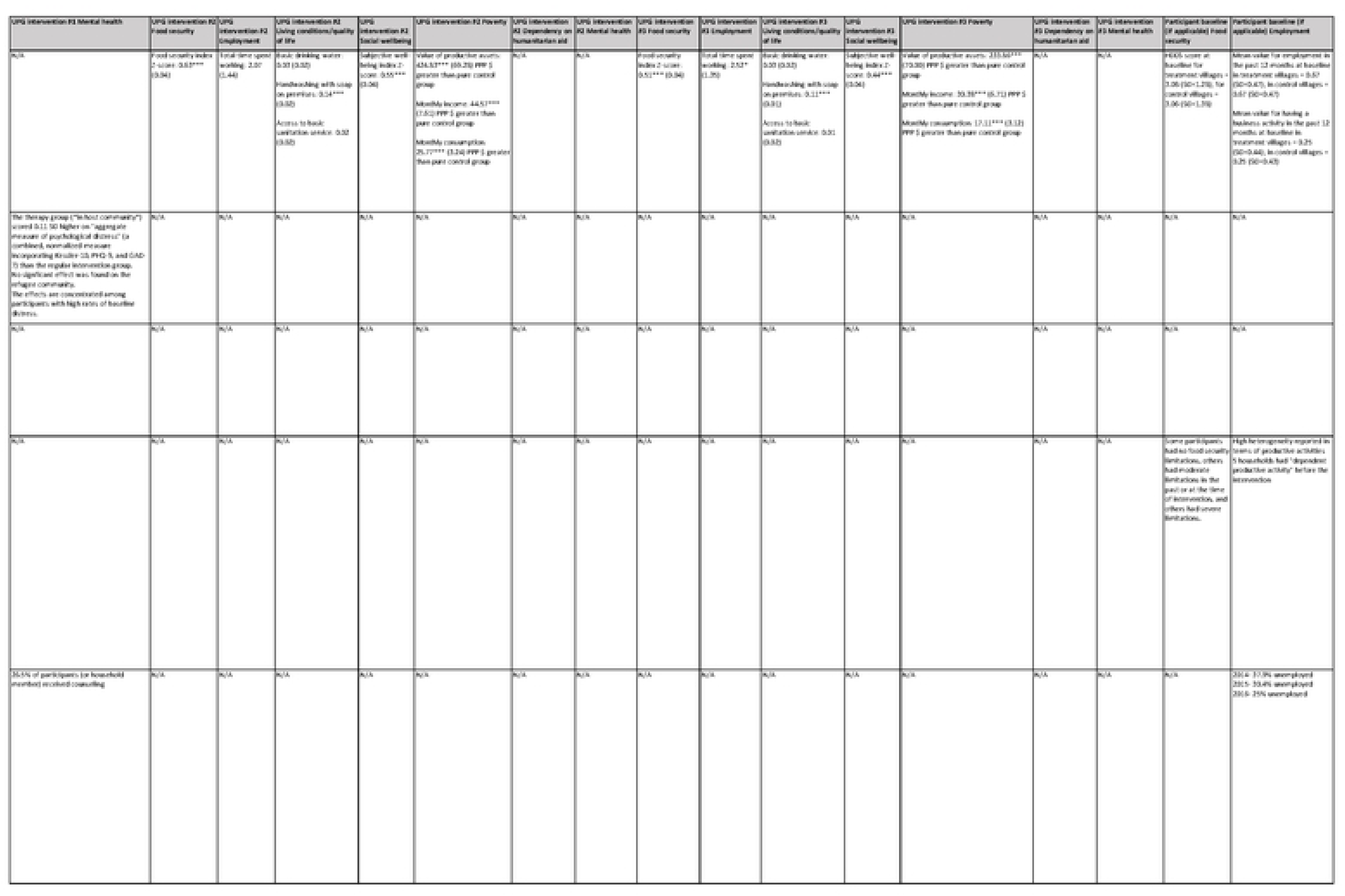
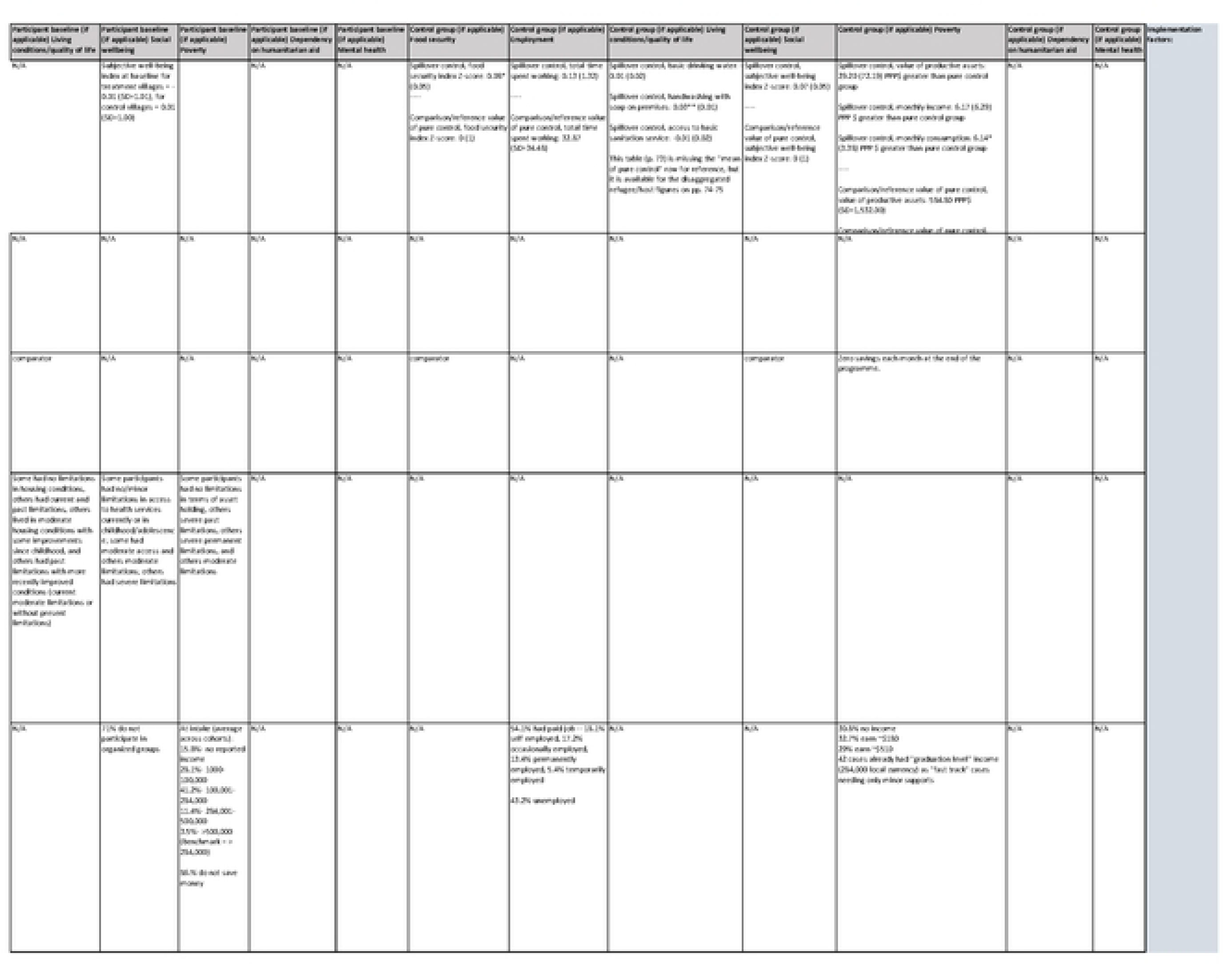
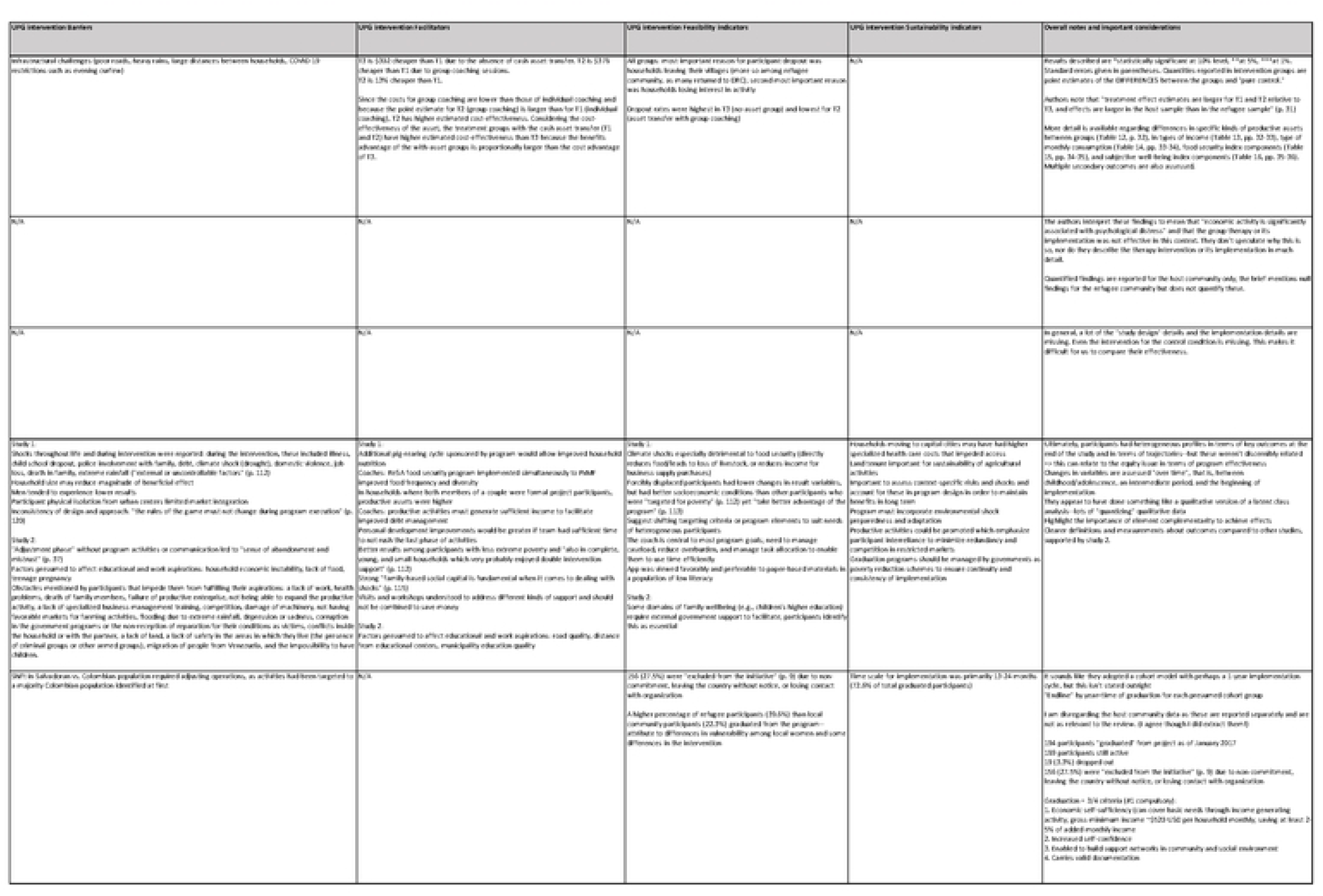
Complete data extraction record.

